# Prostate cancer risk-stratification using proteomic estimation of KLK3 improves accuracy in screening applications: a population-based cohort study in the UK Biobank

**DOI:** 10.1101/2025.08.21.25334151

**Authors:** Anya Morris, Ella Daniels, Jingzhan Lu, Bethan Mallabar-Rimmer, Michael N Weedon, Sarah ER Bailey, Leigh M Jackson, Harry D Green

**Author notes:** corresponding author The Garden Suite, St. Luke’s Campus, Magdalen Road, Exeter, UK.

## Abstract

**Background:** The prostate specific antigen (PSA) test is the most used clinical tool for prostate cancer risk stratification. PSA-based screening remains controversial due to modest predictive power (ROC AUC<0.75), high false-positive rates and racial disparities. Here, we evaluated the prostate cancer risk-stratification potential for the KLK3 protein, measured by Olink® Explore-3072, in the UK Biobank.

**Methods:** 19,364 men in the UK Biobank’s proteomics dataset were cancer-free at assessment centre visit. We used logistic regression to evaluate potential of KLK3 to predict prostate cancer within 2, 5, and 10 years of recording, as an independent predictor and with age and genetic risk score. Prostate cancer cases were classified by severity based on clinical action taken post-diagnosis. All predictive models were performed under 5-fold cross validation, and diagnostic accuracy statistics reported for the test set.

**Findings:** KLK3 was strongly associated with prostate cancer incidence (HR per SD: 3.00 (2.87 - 3.13), p<2e-16). ROC AUC for a 2-year prediction horizon was 0.918 (0.906 - 0.93), reducing to 0.854 (0.848 - 0.86) over 10 years. 10-year ROC AUC was stronger in individuals of European ancestry (0.857 (0.851 - 0.863)) than African (0.801 (0.762 - 0.841)) or South Asian (0.795 (0.727 - 0.862)) ancestry. Power to predict highly aggressive cancer cases within 2 years of recording was strong (ROC AUC 0.928 (0.919 - 0.937)) but weaker for a 10-year period (0.87 (0.865 - 0.876)). Inclusion of age and genetic risk score provided small improvements in individuals of primarily genetic European ancestry, but no improvement in African or South Asian ancestry.

**Interpretation:** The ROCAUC values reported are superior to those seen previously for the PSA test. We demonstrate high predictive accuracy across 2-, 5- and 10-year windows. Findings were consistent for low and high-risk cases. These findings suggest that proteomic PSA measurements may be helpful in prostate cancer risk stratification, while highlighting the need for improved predictive models across diverse ancestral groups.

**Funding:** This study was funded by the University of Exeter. We report no conflicts of interest.

**Research In Context:** *Evidence before this study:* On 4^th^ August 2025 we searched the PubMed database using the search terms [(PSA OR KLK3 OR Prostate Cancer) AND (Risk Prediction OR Screening) OR (UK Biobank AND Proteomics)] to establish predictive power of the PSA test alongside prior work on the UK Biobank Proteomics data and identified a number of relevant studies. Large meta-analyses concerning the PSA test documents moderate predictive accuracy (ROC AUC~0.72), a high false positive rate, and no evidence that PSA screening reduces overall mortality. Studies using the UK Biobank Proteomics dataset have taken a phenome-wide or pan-cancer approach, scanning thousands of potential disease-protein pairs.

*Added value of this study:* We assessed the predictive value of KLK3 (molecularly equivalent to PSA) measured by Olink® high-throughput proteomics in 19,392 cancer-free men from the UK Biobank. We evaluated performance over 2-, 5-, and 10-year prediction horizons and stratified results by ancestry and cancer severity. KLK3 was a strong independent predictor of prostate cancer, particularly in a short window following measurement, and outperformed PSA estimates reported in previous literature. Combining KLK3 with age and a polygenic risk score provided modest benefit in men of European ancestry, but no additional benefit in men of African ancestry. This is the first large-scale study to assess proteomic measurement of KLK3 to stratify prostate cancer risk in a general population cohort.

*Implications of all the available evidence:* Proteomic measurement of KLK3 offers improved risk prediction for prostate cancer compared to standard PSA testing over a 2-, 5-, and 10-year period. Our findings suggest that KLK3 could enhance risk stratification in population screening and may be particularly useful for identifying individuals at very high risk. Similar to current technologies, proteomic measurement of KLK3 performs worse in African and South Asian populations than in European populations. While this disparity highlights an urgent need to improve and validate predictive models in non-European populations, our reported predictive power in non-European populations that is stronger than the current PSA test for European populations. These results suggest that proteomic measurement of KLK3 in screening applications will result in improved accuracy across populations and for severe prostate cancer outcomes over previous technology.

## Introduction

Prostate cancer is the most commonly diagnosed cancer in males in the UK with 55,000 new cases every year, accounting for 28% of new cancer cases in men.^1^ Whilst mortality rates are expected to continue to fall as they have over the last decade, there are still 12,000 deaths per year due to prostate cancer in the UK, with 75% of these occurring in males over the age of 75.^1^ Reduction in mortality rates has been partially attributed to prostate specific antigen (PSA) testing in primary care, and subsequent referral to a specialist for men with elevated PSA levels, which facilitated earlier detection and potentially curative intervention.^2^ However, widespread PSA testing has raised concerns for overdiagnosis, leading to the identification and treatment of clinically indolent tumours that may never have caused clinical symptoms or required treatment.^2^

Lower urinary tract symptoms (LUTS) including increased urinary frequency or urgency, or new onset erectile dysfunction, feature as symptoms of prostate cancer in NICE guidance NG12, which advises GPs on investigating for suspected cancer. In patients with these features, GPs are advised to consider a PSA.^3^ Men over 50 without LUTS can also request a PSA test following a discussion of risks and benefits. Despite being the only routine test for prostate cancer detection, the diagnostic accuracy of PSA is weak and screening is a topic of ongoing debate.^4^

A 2022 meta-analysis of 19 studies found the average receiver operator characteristic (ROC) area under the curve (AUC) for PSA to be 0.72, with a sensitivity and specificity of 0.93 and 0.20 respectively, highlighting the imbalance between detection and false positives.^5^ These findings are consistent with a more recent (2025) meta-analysis concluding PSA testing offers modest mortality benefits, but risks overdiagnosis.^6^ A meta-analysis of five randomised controlled trials enrolling 700,000 men found screening for prostate cancer led to a small reduction in prostate cancer-specific mortality, but no reduction in overall mortality.^7^

Genetic risk scores (GRS), shown to be an independent predictor of prostate cancer, ^8^ While there are concerns that GRS implementation in clinic may exacerbate the overdiagnosis problem,^9^ the 2025 BARCODE1 study found that individuals with a GRS in the top 10% had a higher rate of clinically significant disease (defined by a Gleason score of at least 7) than identified by the PSA test.^10^

The lack of transferability of GRSs across ancestral groups^11^ could exacerbate existing disparities.^12,13^ This is particularly relevant in prostate cancer due to the significantly higher incidence of prostate cancer in Black men compared to White men.^14^ However, while GRSs perform poorly in non-White ancestral groups, the same is true of the PSA test, which is higher in Black men without prostate cancer than in White men without prostate cancer, leading to poor specificity and potentially overdiagnosis.^15^

In 2020, Olink Proteomics launched the UKBiobank Pharma Proteomics Project (UKB-PPP).^16^ This project involved measuring 3000 proteins from blood samples collected at UKB assessment in 55,000 individuals using the Olink Explore 3072 technology. A study predicting 218 diseases using 5-20 proteins identified using a feature-selection strategy found models built using proteomics data outperformed clinical models for 67 pathologically diverse diseases.^17^ A cancer-specific study of 1,463 proteins and 19 cancer sites found three proteins associated with prostate cancer, but KLK3 was not included in the 1,463.^18^

Among these was KLK3 (also known as PSA), encoded by the *KLK3* gene, enabling the assessment of PSA levels in a large, unselected, and population cohort. This dataset provides a unique opportunity to evaluate the predictive utility of PSA outside the context of clinical referrals or opportunistic screening. Hereon, we use ‘PSA’ to refer to the currently used technology in clinic and ‘KLK3’ to refer to the Olink-derived proteomics readings.

The aim of this study was to assess the predictive power of KLK3, measured by Olink proteomics, for incident prostate cancer in the UKBiobank cohort.

## Methods

### Participants

Male UK Biobank (UKB) participants (‘Male’ sex recorded in UK Biobank field 31) of any ancestry that had a KLK3 reading were included in this study. UKB is a cohort study of over 500,000 UK adults aged between 40-70 years at recruitment (2006-2010). The data includes demographics, biomarkers, genetics and linked medical records. For sample size consistency across analyses, we only included participants with linked genetic data and excluded those with prostate cancer prior to the assessment centre visit. For ancestry-specific analyses, genetically inferred ancestry was used. Genetically inferred ancestry was derived from principal component analysis using individuals from the 1000 Genomes Project. The UKB data are available to any bona fide researcher following application. For full information on UKB see Bycroft et al. ^19^

### Phenotype Definition

#### KLK3 protein

KLK3 protein data were obtained from UKB-PPP, which measured ~2,900 proteins from ~50,000 randomly selected UK Biobank participants. ^16^ Protein quantification was performed on blood plasma samples using O-links antibody-based proximity extension assay (PEA), followed by a next generation sequencing (NGS) readout.^20^ Further details on this methodology can be found at https://olink.com/knowledge/documents.

Protein expression levels in UKB are reported in Normalised Protein eXpression (NPX) units, an arbitrary, relative quantification unit on a log2 scale, where a difference of 1 NPX reflects a doubling in protein concentration. ^20^ Absolute quantification, such as pg/ml, is not available so KLK3 values could not be directly compared to GP-recorded PSA values.

#### Prostate cancer

Prostate cancer was defined using ICD10 code C61. Individuals were classed as cases if they had an entry in the linked cancer registry data, hospital episode statistics or death records for this code dated after their assessment date. Anyone who had a prostate cancer diagnosis prior to the assessment date were excluded from analysis.

#### Aggressive prostate cancer

Stage and grade of cancer was not available in the UK Biobank cohort at the time of conducting this study. As a proxy for cancer severity, we used OPCS records and death records to ascertain whether a cancer diagnosis was acted on clinically within 2 years of diagnosis. We classified a prostate cancer diagnosis into three severity groups:

*Severe:* the participant died of any cancer (ICD10 code C* as either primary or contributory cause of death), or undertook chemotherapy (OPCS codes X70, X71 or X72) within 2 years of diagnosis.

*Actionable:* the cancer case results in either radiotherapy (OPCS codes X65, X66, X67 or X69), prostate surgery (OPCS code M61) or anti-androgen treatments (OPCS codes X74.1, X38.3, S52.5 or S52.6) within 2 years of diagnosis, or any of the severe criteria.

*All:* Any record of prostate cancer.

#### Ancestry

We performed subgroup analyses in European (EUR), African (AFR) and South Asian (SAS) genetically inferred ancestry. Principal component analysis was performed using individuals from the 1000 Genomes Project prior to the projection of UK Biobank individuals into the principal component space. K-means clustering was subsequently applied to classify individuals into respective ancestral groups, with centres initiated to the mean principal component values of each 1000 Genomes sub-population.

#### Genetic Risk Score

A genetic risk score for prostate cancer was derived using the 269 risk variants reported in Conti et al.’s^21^ 2021 trans-ancestry genome-wide meta-analysis. Weighting for each single nucleotide polymorphism (SNP) was given by the log of the odds ratio from Supplementary Table 4 of Conti et al. These weights were used over the UK Biobank weights to avoid issues with overfitting. For each UKB participant, the GRS was calculated using the sum of weights multiplied by the participant’s genotype.

### Statistical Methods

All analysis was conducted using R version 4.4.0 “Puppy Cup”. We first evaluated a Cox proportional hazards model to estimate the association between KLK3 levels and 10-year risk of incident prostate cancer. Participants were divided into quintiles based on their KLK3 measurements, with the fifth quintile containing the highest 20% of values. Individuals were followed from the date of assessment to the earliest of prostate cancer diagnosis, death, or 10 years of follow-up. Participants who died before a prostate cancer diagnosis were censored at the date of death. Since recruitment from the UK Biobank was completed in 2010, and the cancer registry linkage is updated to 2021, all participants have the requisite 10 years of follow-up.

Second, we developed an integrated risk model (IRM)^22^ using logistic regression to assess the predictive performance of KLK3 for prostate cancer. We tested KLK3 as an independent predictor, and as part of an integrated risk model alongside age at assessment and prostate cancer GRS. We tested predictive ability for a 2, 5 and 10 year prediction horizon following assessment centre. Anyone cancer-free by the end of the prediction horizon were considered controls.

Predictive performance of the integrated risk model was assessed using a five-fold cross validation with five repeats to reduce overfitting. Discrimination was measured using receiver operator characteristic (ROC) area under the curve (AUC) with 95% confidence intervals (95%CIs). The Youden’s J optimised threshold was identified for each model and used to calculate other diagnostic accuracy statistics such as sensitivity, specificity, positive predictive value (PPV), and negative predictive value (NPV). All diagnostic statistics are reported for the testing dataset.

We repeated our primary analyses excluding any participant who either had a PSA test at any time, or did not have GP record linkage. This sensitivity analysis ensures that predictive power is not affected by a high PSA test causing a diagnosis through differential degrees of clinical follow-up.

### Ethical Approval and Data Availability

Ethics approval for the UK Biobank study was obtained from the North West Centre for Research Ethics Committee (11/NW/0382). Written informed consent was obtained from all participants. All individual-level data used in this paper were obtained from the UK Biobank resource, and can be obtained from the UK Biobank at [https://www.ukbiobank.ac.uk/enable-your-research/apply-for-access]. Information on recruitment, locations and data collection methods can be found in Bycroft et al.^19^.

## Results

### Cohort Description

Of the 502,355 participants in the UKB, 44,792 had KLK3 data, 20,570 of those reported male sex, and 19,670 had linked genetic data. 270 participants were excluded due to a record of prostate cancer prior to the assessment centre visit, leaving a cohort of 19,392. 4.4% of the cohort had a record of prostate cancer within 10 years of the assessment centre visit. Prostate cancer incidence was higher in participants of AFR ancestry than those of EUR ancestry, but total numbers were very low for 2 and 5 years.

Of the 859 cases within 10 years of the assessment centre, 304 (35.4%) participants required either radiotherapy or surgery within the 2 years following diagnosis (actionable), and 86 (10%) of cases either died of a cancer-related death or undertook chemotherapy within 2 years (severe).

Due to the low numbers of aggressive cancer cases and cases in AFR and SAS ancestry, we only report diagnostic accuracy statistics for these subgroups for the 10-year prediction horizon.

### Risk stratification ability of KLK3

Circulating KLK3 levels associated strongly with prostate cancer incidence. In a Cox-PH model, one standard deviation increase in KLK3 associated with a 3 fold increase in hazard (95% CI: 2.87 - 3.13, p<2e-16). As shown in Figure 2, this association is highly nonlinear, with minimal risk in the lowest quintiles: 10-year cancer risk was under 1% in quintiles 1 and 2, 1.18% (95% CI: 1.52 - 0.83) in quintile 3, and 3.5% (95% CI: 4.08 - 2.91) in quintile 4. Individuals in the top 20% of the KLK3 distribution had a 10-year cancer risk of 17.18% (95% CI: 18.39 - 15.95), equivalent to a 13.1-fold increase in risk for compared to the rest of the cohort (95% CI: 11.2 – 15.3, p<2e-16).

**Figure 1.**
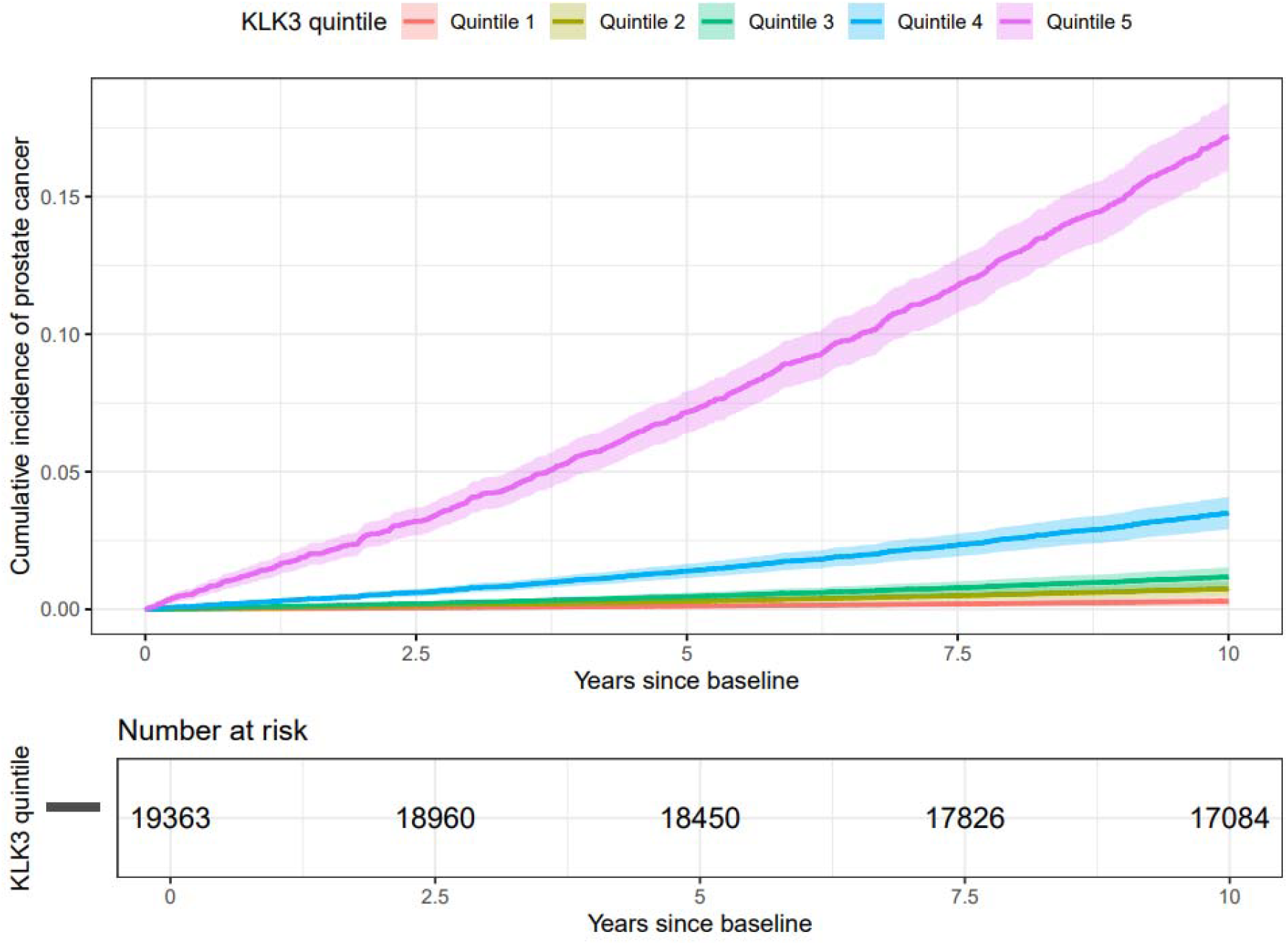
Cumulative incidence of prostate cancer by KLK3 quintile. Prostate cancer incidence was significantly higher in individuals with higher KLK3 levels at the time of UK Biobank assessment centre visit. Individuals in the top quintile had a risk 13 times higher than the rest of the cohort.

**Figure 2.**
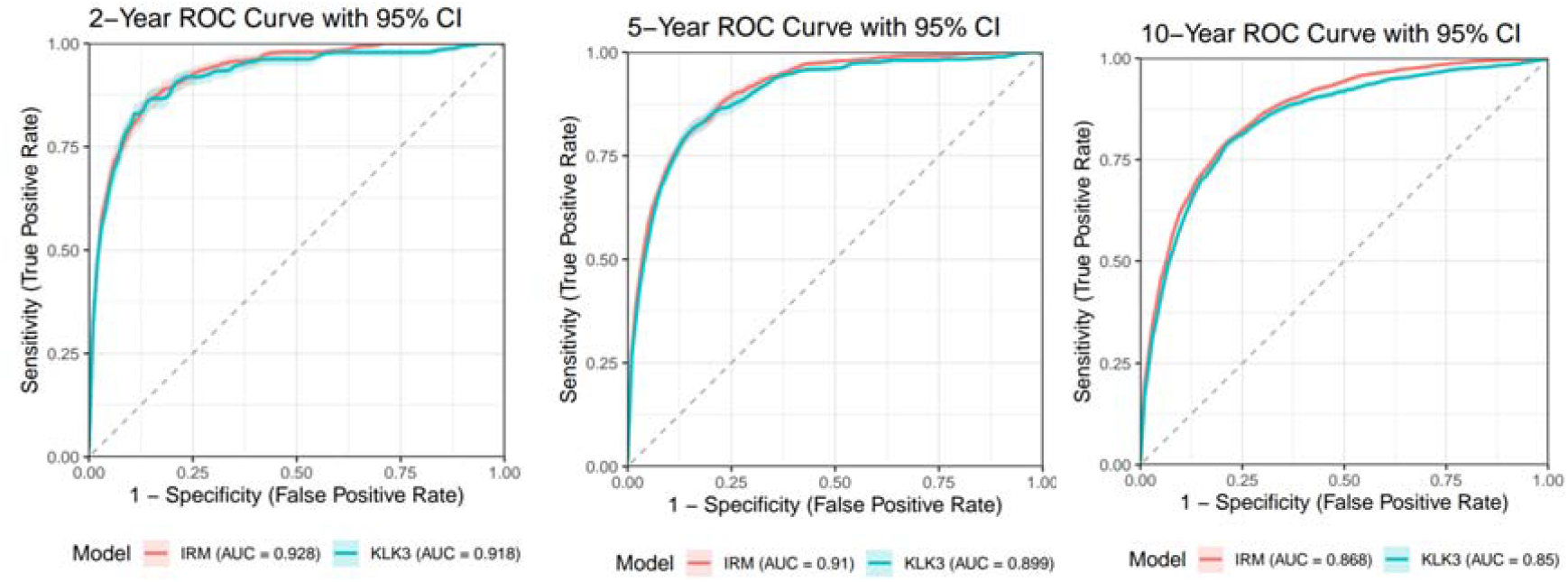
ROC curves by prediction horizon and model type. For all prediction horizons, KLK3 as an independent predictor had a similar ROC AUC to an integrated risk model (IRM). For both models, predictive accuracy dropped as prediction horizon lengthened.

**Figure 3.**
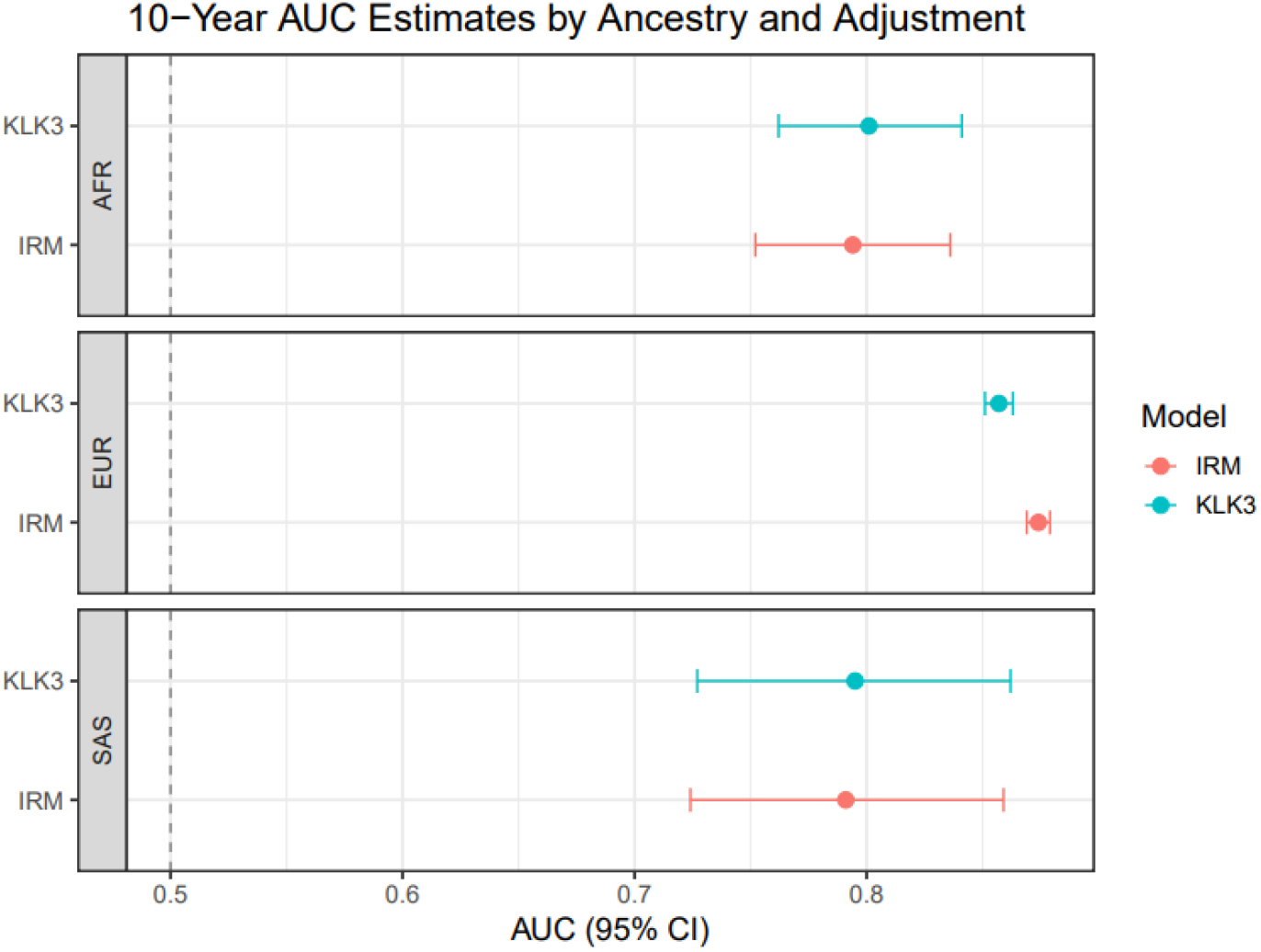
Model Performance by Ancestry. This forest plot shows the ROC AUC and 95% confidence interval for a 10 year prediction horizon, stratified by ancestral group. Performance was lower in individuals of AFR and SAS ancestry compared to individuals of EUR ancestry. Including genetic risk score and age in an integrated risk model improved performance in EUR ancestry, but not in AFR or SAS ancestry.

In a logistic regression model, circulating KLK3 levels showed strong power to predict prostate cancer. The ROC AUC for predicting prostate cancer within 2 years of the assessment centre was 0.918 (0.906 - 0.93), with a sensitivity and specificity both above 0.8. The ROC AUC reduced to 0.854 (0.848 - 0.86) when predicting prostate cancer within 10 years of the assessment centre.

A longer prediction horizon associated with a reduction in ROC AUC, Sensitivity, Specificity and NPV, but an increase in PPV (Table1) and due to increased case/control ratio the further the prediction horizon is extended. In individuals with linked GP records who had never had a PSA test, the predictive power of KLK3 remained strong (2-year ROC AUC 0.893 (0.866 - 0.919) and 10-year ROC AUC 0.865 (0.855 - 0.875)).

**Table 1.**
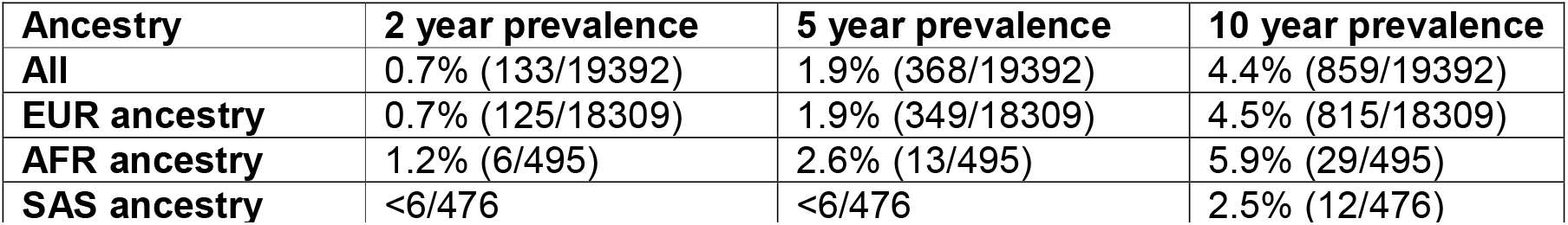
Cohort description. This table shows the breakdown of the 19,392 individuals eligible for this study. Data are formatted as % (cases/total). 94.4% of the participants included were of EUR ancestry, 2.6% were of AFR ancestry, 2.5% were of SAS ancestry and 0.5% were of other ancestral groups. Prostate cancer prevalence was roughly 50% higher in the AFR participants than in EUR participants. AFR: African ancestry. EUR: European ancestry. SAS: South Asian ancestry. Numbers <6 have been omitted due to the UK Biobank’s guidelines on reporting low numbers.

### An integrated risk model

The inclusion of age and genetic risk score in an integrated risk model did not significantly increase the performance of the model with a 2 year prediction horizon (0.928 (0.919 - 0.937) vs 0.918 (0.906 - 0.93)). However, for a 10 year prediction horizon, the integrated risk model provided a small but statistically significant increase in ROC AUC (0.854 (0.848 - 0.86) vs 0.87 (0.865 - 0.876)). Full diagnostic statistics for the integrated risk model can be found in supplementary table 2.

**Table 2.** Diagnostic Statistics of KLK3 in the UK Biobank. Data are reported statistic (95% CI). All statistics are reported from the testing set of a 5-fold cross validated logistic regression model optimised by Youden’s J statistic. Prediction Horizon represents the time window from which cases were defined, starting from the assessment centre visit. ROC AUC (receiver operating characteristic area under curve), PPV (positive predictive value), NPV (negative predictive value).

| Prediction Horizon | 2 years | 5 years | 10 years |
| --- | --- | --- | --- |
| ROC AUC | 0.918 (0.906 - 0.93) | 0.903 (0.896 - 0.91) | 0.854 (0.848 - 0.86) |
| Sensitivity | 0.862 (0.833 - 0.887) | 0.826 (0.807 - 0.843) | 0.788 (0.776 - 0.8) |
| Specificity | 0.859 (0.856 - 0.861) | 0.838 (0.836 - 0.841) | 0.791 (0.788 - 0.794) |
| PPV | 0.0404 (0.0373 - 0.0438) | 0.09 (0.0857 - 0.0944) | 0.149 (0.144 - 0.154) |
| NPV | 0.999 (0.999 - 0.999) | 0.996 (0.996 - 0.996) | 0.988 (0.987 - 0.989) |

### Predictive performance by ancestral group

KLK3 performed better as a predictor in individuals of EUR ancestry (ROC AUC 0.857 (0.851 - 0.863)) vs AFR (ROC AUC 0.801 (0.762 - 0.841)) or SAS (ROC AUC 0.795 (0.727 - 0.862)) ancestry. This disparity was exacerbated when GRS and age were added to an IRM, as doing so significantly increased the predictive power in EUR ancestry (ROC AUC 0.874 (0.869 - 0.879)) but provided no benefit in AFR ancestry (ROC AUC 0.794 (0.752 - 0.836)) or SAS ancestry (0.791 (0.724 - 0.859). Full diagnostic statistics for 2, 5 and 10 year prediction horizons can be found in supplementary tables 3-4 (EUR Ancestry), 5-6 (AFR ancestry) and 7-8 (SAS ancestry).

### Aggressive Cancer Outcomes

For up to a 5 year prediction horizon, ROC AUC for KLK3 as an independent predictor remained similar for all cases (0.903 (0.896 - 0.91)), actionable cases (0.905 (0.895 - 0.916)) and severe cases (0.901 (0.872 - 0.931)). However, when the prediction horizon was extended to 10 years, KLK3 performed significantly better for all (0.854 (0.848 - 0.86)) and actionable cases (0.843 (0.833 - 0.853)) compared to the most severe cases (0.772 (0.745 - 0.798)). Similar trends are observable in the IRM, which performed better to predict 10-year severe outcomes (0.8 (0.778 - 0.822)) (Figure 4). Full diagnostic accuracy statistics are available in Supplementary Tables 9-10 (actionable) and 11-12 (severe).

**Figure 4.**
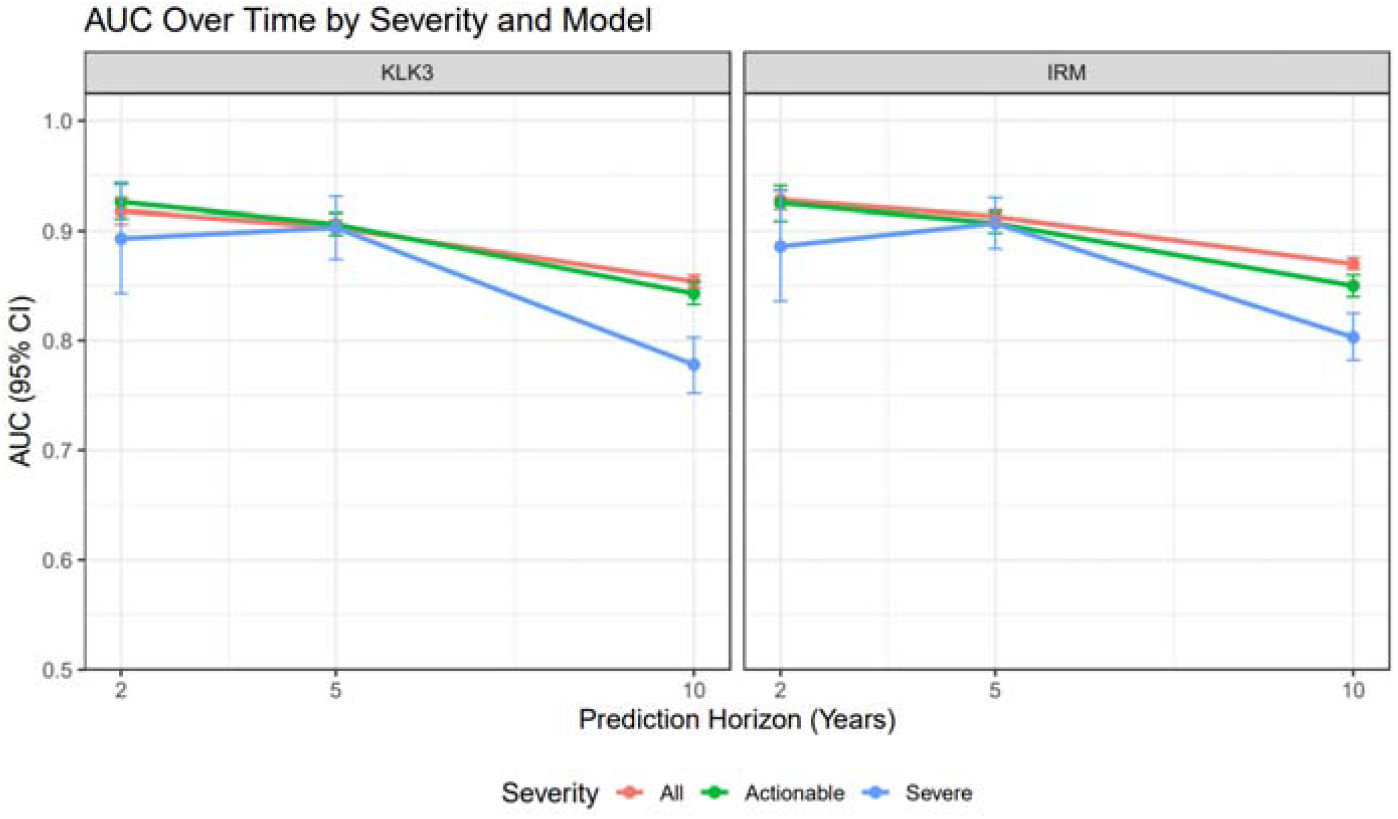
Model performance by cancer aggressiveness. A line graph showing ROC AUCs and 95% confidence intervals for 2, 5 and 10 year prediction horizons for all cancer cases (red), actionable prostate cancer (green) and severe cancer cases (blue).Performance was similar for all, actionable and severe cases up to 5 years, but for both KLK3 as an independent predictor and as part of an integrated risk model, performance for severe prostate cancer cases was weaker for a 10 year prediction horizon.

## Discussion

### Summary of Results

Our results demonstrate the predictive power of proteomic measurement of KLK3 for incident prostate cancer in the UKB population cohort. We have shown that KLK3 is strongly predictive of prostate cancer up to a 10 year prediction horizon. KLK3 was a strong predictive marker of actionable and severe disease up to 5 years, but declined over a 10 year period, and was lower in individuals of AFR and SAS ancestry. This pattern of results is consistent with existing evidence from PSA-based screening studies and smaller clinical studies showing clinical utility of proteomics analysis in cancer.^23^

Inclusion of genetic risk and age offered minor improvements to predictive performance, most notably in long-term prediction of severe cases, but did not improve performance in African or South Asian ancestry. Lower performance of genetic risk scores in non-White ethnicities is a well-documented issue in genomics studies,^24^ and although a trans-ancestral GRS was used,^21^ the GRS weights were still based on an 80% European population. The discrepancy in both KLK3 performance and in value added by a GRS highlight the importance of addressing racial disparities in genomic and proteomic research.

Our observed predictive performance (ROC AUC 0.918 at 2 years and 0.854 at 10 years) substantially exceeds the accuracy typically seen in conventional PSA based screening (ROC AUC < 0.75).^5^ Although we report lower predictive performance in AFR ancestry (0.801), SAS ancestry (0.795) and for 10-year prediction of severe cancer cases (0.772), the values we observe are still stronger than current PSA testing technologies.

### Strengths and Limitations

A major strength of our study is the scale of the data available in the UK Biobank and the duration of follow-up. Due to the prospective nature of the recruitment, our sample reflects a real-world general population, and has not been selected based on clinical features. Recruitment between 40 and 70 from 2006 to 2010 covers the peak incidence period of prostate cancer and provides at least 10 years of follow-up data from cancer registry and hospital episode statistics. As the Olink Explore 3072 platform was first formally announced in 2021, it is not possible to perform such a study and have 10 years of follow-up data in any other cohort. Where previous studies analysed the proteomics data under a wide scope, scanning multiple conditions with thousands of proteins,^17,18^ our study focuses on evaluating the diagnostic potential of KLK3 as a clinical tool, considering ancestry, severity and clinically relevant diagnostic statistics.

However, we acknowledge several limitations. The UK Biobank cohort has a well-known bias towards healthy individuals at time of recruitment. Despite the Biobank’s large sample size, the number of men of AFR or SAS ancestry with linked proteomics data led to imprecise ROC AUC estimates, particularly for short time prediction horizons. Since the UK Biobank’s cancer registry data does not have stage and grade of cancer, we relied on clinical action as a proxy. Future prospective studies using Gleason scores to define clinically significant cancers (like in BARCODE1) are necessary. A consequence of the UK Biobank being the only cohort in which this research is currently possible is that no independent validation cohort exists, and we used a cross-validation approach to controlling for overfitting. However, a case-control study of ~40,000 Chinese participants with 5.7 years of follow-up found similar results when using UKB-PPP for replication.^25^

There are also limitations in interpretation of results. The proteomic assay used provides relative, not absolute, KLK3 quantification, which is not possible to convert into a value comparable with the values from the PSA test.

At the time of conducting this study, clinical implementation would be inhibited by cost. Rates quoted for the Olink Explore 3072 cover a wide range from £150 - £600. The targeted protein panels are typically much cheaper (~£30) but current Olink panels do not contain KLK3 and results would not be immediately transferable from the UKB-PPP data. The PSA test costs ~£21 per test according to a 2022 cost-effectiveness analysis, but follow-up tests include MRI (£339), biopsy (£581).^26^ For a medicine to be considered cost-effective, NICE guidelines require no more than £20,000 - £30,000 per quality-adjusted life years gained. A comprehensive health economics analysis is necessary to determine if the diagnostic improvements from KLK3 would be cost-effective.

### Implications

Our results have important implications for clinical practice, public health policy and prostate cancer research. The superior short-term predictive accuracy of proteomic KLK3 measurement suggests a role for high-throughput proteomics in targeted prostate cancer screening programs over conventional PSA testing. The improved accuracy from this technology has the potential to reduce false positive rates, thereby reducing harm from unnecessary invasive examination. The clear ancestry-based differences in performance highlight an urgent need to develop and validate ancestry-inclusive predictive models.

## Data Availability

All individual-level data used in this paper were obtained from the UK Biobank resource, and can be obtained from the UK Biobank at [https://www.ukbiobank.ac.uk/enable-your-research/apply-for-access]. Information on recruitment, locations and data collection methods can be found in Bycroft et al.19.

## Acknowledgements

This study was supported by the National Institute for Health and Care Research Exeter Biomedical Research Centre. The views expressed are those of the author(s) and not necessarily those of the NIHR or the Department of Health and Social Care. This study has been conducted using the UK Biobank Resource (Application No. 103356).

## Author Contributions (CRediT taxonomy)

Anya Morris: Data Curation, Statistical Analysis, Writing – Original Draft Preparation, Writing – Review and Editing

Jingzhan Lu: Data Curation, Writing – Review and Editing

Ella Daniels: Writing – Review and Editing

Bethan Mallabar-Rimmer: Conceptualisation, Writing – Review and Editing

\Michael Weedon: Writing – Review and Editing

Sarah Bailey – Conceptualisation, Writing – Review and Editing

Leigh Jackson – Statistical Analysis, Writing - Review and Editing

Harry Green – Conceptualisation, Data Curation, Statistical Analysis, Writing – Original Draft Preparation, Writing – Review and Editing

## Supplementary Information

**Supplementary Table 1** – Full diagnostic statistics from KLK3 as an independent predictor in the UK Biobank cohort to predict any prostate cancer case over 2, 5 and 10 year periods.

**Supplementary Table 2** – Full diagnostic statistics from an integrated risk model including KLK3, age and genetic risk score in the UK Biobank cohort to predict any prostate cancer case over 2, 5 and 10 year periods.

**Supplementary Table 3** – Full diagnostic statistics from KLK3 as an independent predictor in UK Biobank cohort participants of European ancestry to predict any prostate cancer case over 2, 5 and 10 year periods.

**Supplementary Table 4** – Full diagnostic statistics from an integrated risk model including KLK3, age and genetic risk score in the UK Biobank cohort participants of European ancestry to predict any prostate cancer case over 2, 5 and 10 year periods.

**Supplementary Table 5** – Full diagnostic statistics from KLK3 as an independent predictor in the UK Biobank cohort participants of African ancestry to predict any prostate cancer case over 2, 5 and 10 year periods.

**Supplementary Table 6** – Full diagnostic statistics from an integrated risk model including KLK3, age and genetic risk score in the UK Biobank cohort participants of African ancestry to predict any prostate cancer case over 2, 5 and 10 year periods.

**Supplementary Table 7** – Full diagnostic statistics from KLK3 as an independent predictor in the UK Biobank cohort participants of South Asian ancestry to predict any prostate cancer case over 2, 5 and 10 year periods.

**Supplementary Table 8** – Full diagnostic statistics from an integrated risk model including KLK3, age and genetic risk score in the UK Biobank cohort participants of South Asian ancestry to predict any prostate cancer case over 2, 5 and 10 year periods.

**Supplementary Table 9** – Full diagnostic statistics from KLK3 as an independent predictor in the UK Biobank cohort to predict clinical actionable prostate cancer over 2, 5 and 10 year periods.

**Supplementary Table 10** – Full diagnostic statistics from an integrated risk model including KLK3, age and genetic risk score in the UK Biobank cohort to predict clinical actionable prostate cancer over 2, 5 and 10 year periods.

**Supplementary Table 11** – Full diagnostic statistics from KLK3 as an independent predictor in the UK Biobank cohort to predict clinically aggressive prostate cancer over 2, 5 and 10 year periods.

**Supplementary Table 12** – Full diagnostic statistics from an integrated risk model including KLK3, age and genetic risk score in the UK Biobank cohort to predict clinically aggressive prostate cancer over 2, 5 and 10 year periods.

**Supplementary Table 13** – Full diagnostic statistics from KLK3 as an independent predictor in only individuals with linked GP record data but no PSA test recorded in the UK Biobank cohort to predict any prostate cancer case over 2, 5 and 10 year periods.

